# Trial of an individualised Intervention for the prevention of Stroke (TIIPS): Protocol and statistical analysis plan for a randomized controlled trial

**DOI:** 10.64898/2026.05.07.26352701

**Authors:** Valery Feigin, Rita V Krishnamurthi, Devaki DeSilva, Lily Yee, Jesse Dyer, Bala Nair, Braden Te Ao, Derrick Bennett, Bruce Arroll, Lillian Choy, Yogini Rathnasabapathy, Suzanne Barker-Collo, Irene Zeng

## Abstract

**Rationale:** Up to 90% of strokes are preventable through the modification and control of lifestyle risk factors. Health and Wellness (HWC) coaching is an established psychological intervention that may address multiple risk factors, including high blood pressure to reduce the risk of stroke.

**Aims:** To determine the effectiveness of HWC in the management of blood pressure and stroke-related modifiable risk factors in reducing the risk of stroke.

**Methods:** This Phase III, open-label, single-blinded, two-arm randomised controlled trial recruited adults with first-ever or recurrent minor stroke or transient ischaemic attack from hospitals in Auckland and Hamilton, New Zealand. Eligible participants were ≥18 years, independent in activities of daily living, had at least two modifiable cardiovascular risk factors, elevated or treated systolic blood pressure, were English-speaking, and had no history of major stroke, myocardial infarction, significant cognitive or mood disorders, or terminal illness. Longitudinal outcomes will be analysed using linear mixed-effects models under an intention-to-treat framework, with time-to-event outcomes analysed using competing-risk methods and missing data handled using multiple imputation with pooling based on Rubin’s rules.

**Study outcomes:** The primary outcome is difference in the mean change from baseline systolic blood pressure (SBP) to 6-months post-randomisation between control (Usual Care, UC) and HWC groups. The study (n=360) is powered 85% (two sided α=0.05) to detect a mean difference in change of SBP 6 mm Hg (SD ± 20 mm Hg) between HWC and UC groups at 6-months post-randomisation, accounting for a 20% attrition rate. A revised sample size calculation due to a lower attrition rate (9%) provided a required sample size of 320. Secondary outcomes include cardiovascular health score using the Life’s Simple 7; stroke awareness; quality of life; satisfaction with life: cognition; mood; medication adherence; adverse cardiovascular events; health and service costs and productivity status.

**Discussion:** HWC has the potential to modify lifestyle risk factors for stroke. This trial will be the first to test the effectiveness of HWC to modify lifestyle risk factors for secondary stroke prevention.

**Ethical approval:** The trial was approved by the New Zealand Health and Disability Ethics Committee (#2022 EXP 124562022), and The Auckland University of Technology Ethics Committee (#22/206).

**Trial registration:** ACTRN12622000939796 (registered 01/07/2022)

## Introduction

Patients with transient ischaemic attack (TIA) and minor (non-disabling; National Institutes of Health Stroke Scale (NIHSS) ≤ 3)^1^ stroke are at high risk of secondary vascular events, including major stroke, myocardial infarction (MI), cognitive deficits, and death, with population-based studies reporting incidence of adverse outcomes of up to 51.3%^2^. New vascular events, including fatal strokes, MI, and other cardiovascular deaths, occur in up to 26% of patients within four years post-TIA.^3,4^ Increased risk is associated with inadequate secondary stroke prevention management, including unhealthy lifestyle and poor adherence to prescribed medications for the treatment of elevated blood pressure, diabetes mellitus, and previous vascular disease.^3^ The age standardised incidence of first-ever TIA in New Zealand (NZ) is one of the highest among developed countries at 50 [95%CI 46-55] cases per 100,000 persons in 2011-2012.^5^ TIA occurs at a younger mean age in Māori and Pacific people (60 years), and Asian and other (including Middle Eastern and African) people (68 years), compared to Europeans (74 years).^5^ The most recent ARCOS-V also found a high prevalence of cardiometabolic risk factors (e.g., 67% had hypertension, 43% had elevated LDL-cholesterol, and 20% had atrial fibrillation).^6^

### Secondary prevention after TIA/minor stroke

There is evidence that controlling metabolic risk factors and modifying health behaviours for stroke and cardiovascular disease (CVD) prevention is feasible, improves health outcomes, reduces healthcare costs, can reduce individual risk of stroke by about 80%,^7,8^ and can reduce stroke incidence by about 50%.^9^ Addressing health behaviours, including use of multifactorial lifestyle interventions,^10^ may lead to clinically meaningful reductions in CVD and stroke.^11^ Both TIA and minor stroke are highly preventable with medical management,^12-14^ combined with education about stroke/TIA, improved medication adherence, and support for lifestyle behaviour change.^15-18^ Low adherence to healthy lifestyle and prescribed medications, lead to preventable major secondary events.^19-21^

### Health and Wellness Coaching

Well-designed health coaching interventions improve physical and mental health, and sustain changes in lifestyle-related behaviours in people with diabetes,^22,23^ myocardial infarction (MI),^24^ and other chronic conditions.^103^ Resultant health behaviour changes have the potential to be long-lasting.^25^

Health and Wellness coaching (HWC) is a multidimensional psychological behaviour change intervention aimed at improving self-management of lifestyle behaviour and maintaining health and wellbeing.^26^ HWC is a goal-oriented, theory-based,^27^ person-centred partnership that has produced positive effects on health and enhanced well-being of patients with chronic disease.^28-30^ HWC is a widely accepted and established intervention in the community,^31^ and is of particular relevance to stroke prevention as it can address multiple risk factors. HWC fosters ongoing self-directed learning,^28^ delivers a cost-effective^32^ intervention in person or by telephone, and by medical or non-medical personnel, thus reducing cost and increasing the scope of implementation. Individuals who receive HWC have increased perceived health status, improved medication adherence, and physical activity,^33,34^ and significantly improved health outcomes following MI.^35-37^

### Methodological Considerations

A recent systematic review including 15 trials on lifestyle interventions for secondary prevention following TIA or ischemic stroke, with the majority based on educational material, lifestyle advice, or exercise training,^38^ showed a significant lowering of systolic blood pressure (3.6 mm Hg [95% CI -6 to -5.6-1.6]), but no significant effect on cholesterol or mortality. The authors recommended that future trials test interventions with at least eight contact points, using a theoretical framework,^27^ including educational and behavioural interventions with at least a four-month follow-up, and considering factors such as self-efficacy to facilitate health behaviour change.^38^

Testing an intervention that targets brain and heart health requires an evidence-based, relevant and reliable measure to determine its efficacy. The INTERSTROKE study found that ten potentially modifiable risk factors are collectively associated with about 90% of the population-attributable risk of stroke,^39^ including lifestyle related risk factors such as physical activity, diet, and smoking. Of these, hypertension or high blood pressure is the single most significant stroke risk factor.^39^ A meta-analysis of 48 randomised trials evaluating the effects of blood pressure lowering treatments on the risk of major CVD events (including stroke) found that a reduction of 5 mm Hg systolic blood pressure was associated with an 11% reduction in major cardiovascular events in people with previous cardiovascular disease.^40^

Measurement of lifestyle and metabolic risks is also essential to capture the impact of the intervention of health behaviour. The Life’s Simple 7 (LS7) was developed by the American Heart Association (AHA) to predict ideal cardiovascular health using seven domains or metrics,^41-43^ including blood pressure, cholesterol, glucose, body mass index, smoking, physical activity, and diet.^42^

While reducing secondary stroke is the overall aim, the incidence of secondary stroke and TIA events in this population is 11%.^6,44^ Thus, capturing a sufficient number of events would require long-term follow-up and a prohibitively large sample size.^18,38,45^ BP lowering is a good surrogate endpoint for CVD events for preventing CVD and improving total health. ^40^

## Methods

### Ethics and Registration

We conducted cultural consultation with institutional Māori and Pacific advisors prior to study commencement and ethical approval. Ethical approvals from NZ Health and Disability Ethic Committee (AM 12456) and Auckland University of Technology Ethics Committee (22/206). The trial is registered at the Australia New Zealand Trial Registry # ACTRN12622000939796 (registered 01/07/2022).

### Study design

The Trial of an Individualised Intervention for the Prevention of Stroke (TIIPS) is a phase III, prospective, open-label, single-blinded endpoint randomised controlled trial (see Figure 1 for overall trial design). The study will recruit stroke and TIA patients from public hospitals, including outpatient TIA clinics, in Auckland, New Zealand. Recruitment from the existing health system will maximise the intervention uptake. The participating hospitals including Auckland City, Middlemore, North Shore and Waitakere Hospitals in Auckland, and Waikato Hospital in Hamilton.

**Figure 1.**
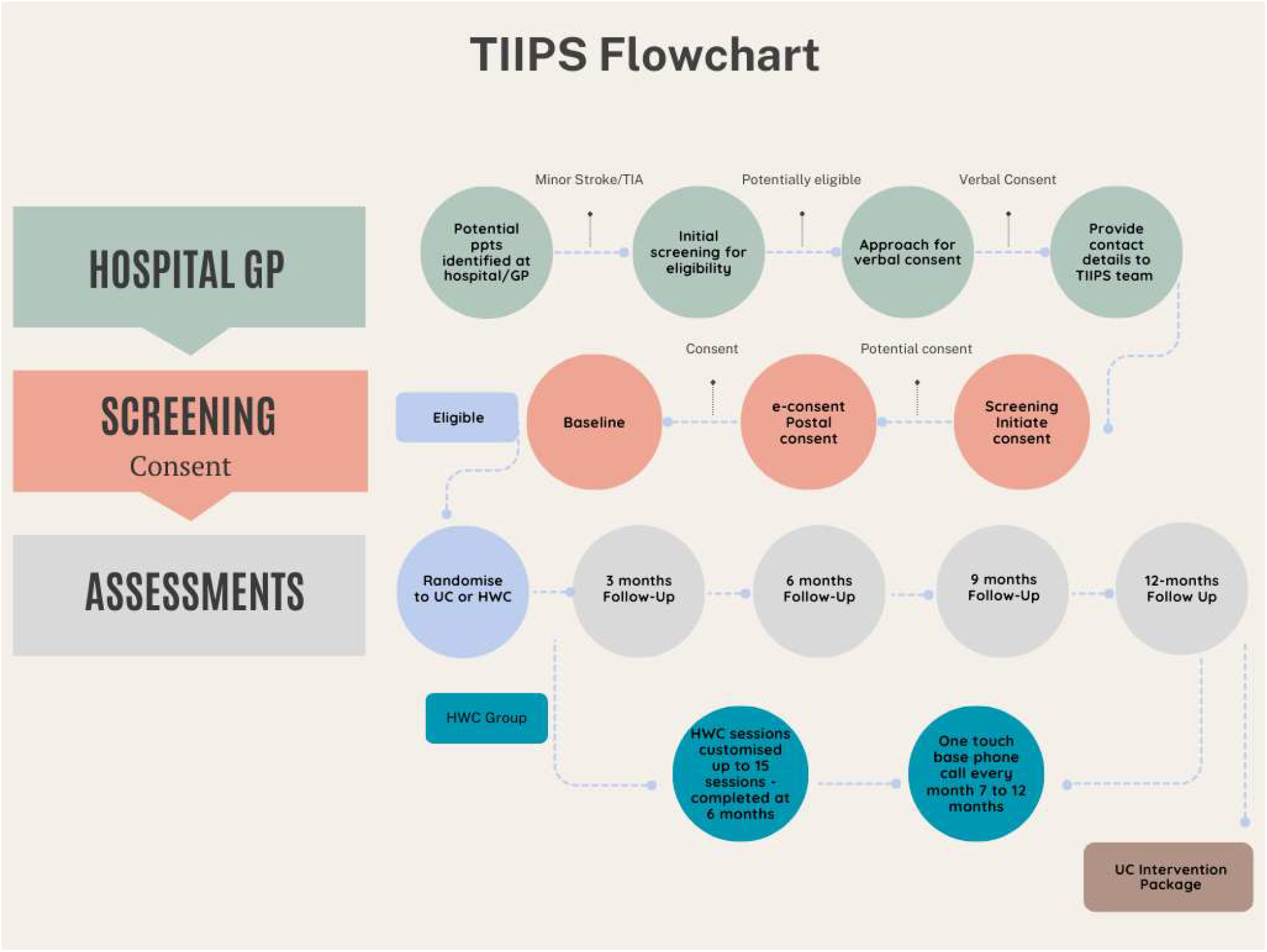
Flowchart of TIIPS study processes.

### Patient population

#### Inclusion criteria

People aged 18 years or older; diagnosed with TIA or minor ischemic or intracerebral haemorrhage stroke (NIHSS ≤ 4) and/or the modified Rankin Scale (mRS) score 0-2 at discharge^1^ or independent in activities of daily living in the past 90 days; admitted to one of the five identified hospitals, or identified via primary care for minor stroke or TIA; with at least 2 modifiable risk factors; who can converse in English; and provide written informed consent.

#### Exclusion criteria

History of major stroke or MI (verified through medical records); subarachnoid haemorrhage; planned carotid endarterectomy; life-threatening conditions with a life-expectancy <5 years; significant clinical depression/anxiety, either in clinical records in the past year or at study screening (Hospital Anxiety and Depression Scale (HADS) score ≥ 11 for either depression and/or anxiety domains); diagnosis of psychiatric conditions in clinical records; history (past year) of alcohol or drug/substance abuse (based on medical records); dependent on others (living in a rest-home/care facility); pre-existing diagnosis of dementia; cognitive impairment at screening (MoCA <23); participation in another RCT.

Exclusion due to significant anxiety and depression or cognitive impairment is necessary in order to recruit participants who are able to engage effectively with the study over a period of 12-months.

The trial protocol will be reported in accordance with the SPIRIT (Standard Protocol Items: Recommendations for Interventional Trials) statement^46^. The study will be reported in accordance with Consolidated Standards of Reporting Trials (CONSORT)^47,48^ (Figure 2), and the Template for Intervention Description and Replication (TiDIER) checklist.^49^

**Figure 2.**
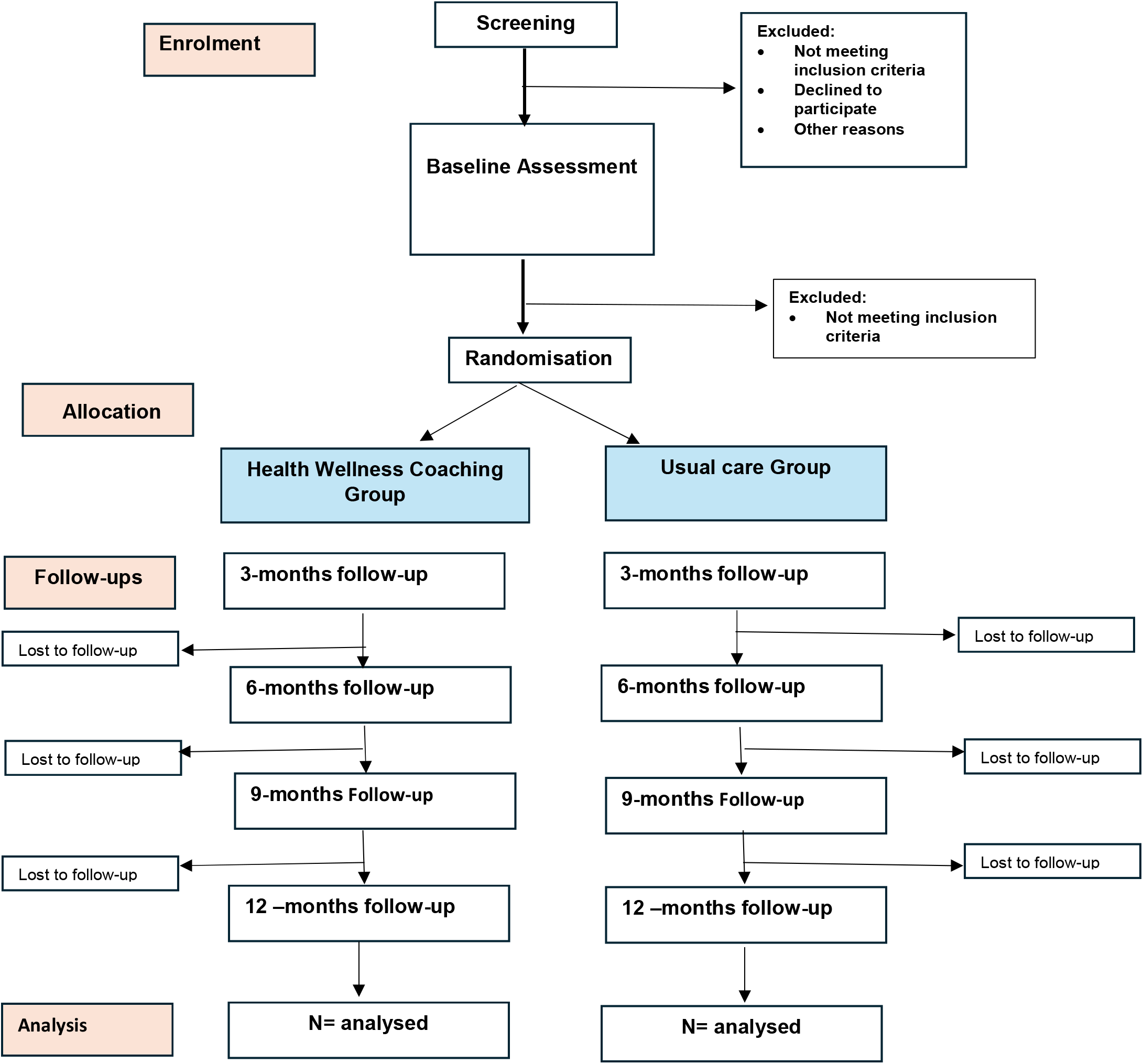
CONSORT Flow Diagram

### Recruitment and randomization

#### Screening

Patients admitted to the recruitment sites with a diagnosis of ischaemic stroke, intracerebral haemorrhage or TIA as confirmed by their treating physician and CT/MRI brain scanning on admission will be screened by a hospital-based research nurse for the main inclusion and exclusion criteria. Those who meet the criteria will be approached for verbal consent to be contacted by a study research assistant (RA) for further information (see Table 1) and written informed consent. The number of people who meet the initial screening criteria will be registered in the study database. Participants who meet all the study criteria will complete a baseline assessment and proceed to randomisation. Criteria not met will be recorded for those who are not included in the trial.

**Table 1.** Demographic and Outcome Assessments.

|  | Baseline<sup>a</sup> | 3<sup>b</sup> | 6<sup>a</sup> | 9<sup>b</sup> | 12<sup>b</sup> |
| --- | --- | --- | --- | --- | --- |
| Demographic Factors: Age, sex, ethnicity, employment, education, marital status | ✓ |  |  |  |  |
| Event type - stroke and pathological subtypes, or TIA and event date (physician diagnosed) | ✓ |  |  |  |  |
| NIHSS or mRS | ✓ |  |  |  |  |
| Hospitalization details: hospital, date of admission, date of discharge | ✓ |  |  |  |  |
| Event details (revascularization and planned procedures) from medical records | ✓ |  |  |  |  |
| Hospital Anxiety and Depression Scale (HADS) | ✓ | ✓ | ✓ | ✓ | ✓ |
| Stroke awareness (recognition of risk factors, knowledge of actions) | ✓ |  | ✓ |  | ✓ |
| Quality of life (EQ-5D-5L) | ✓ |  | ✓ |  |  |
| Life Satisfaction Sliding Scale | ✓ | ✓ | ✓ | ✓ | ✓ |
| Satisfaction with life scale | ✓ | ✓ | ✓ | ✓ | ✓ |
| Cognitive functioning (MoCA) | ✓ |  | ✓ |  |  |
| Medication adherence and self-efficacy (SEAMS) | ✓ | ✓ | ✓ | ✓ | ✓ |
| Physical measurements (non-fasting blood test, SBP/DBP, BMI, HR)* | ✓ |  | ✓ |  |  |
| Absolute and relative 5-year risk of stroke (as measured by Stroke Riskometer app) | ✓ |  | ✓ |  |  |
| Lifestyle factors (Diet score, physical activity, smoking, alcohol) (LS7) | ✓ | ✓ | ✓ | ✓ | ✓ |
| CVD outcomes, recurrent events, hospitalisation (stroke, CVD events) self-report and/or from clinical records | ✓ | ✓ | ✓ | ✓ | ✓ |
| Health and Service costs: New Zealand National Minimum Dataset |  | ✓ | ✓ | ✓ | ✓ |
| Productivity level: New Zealand National Minimum Dataset |  | ✓ | ✓ | ✓ | ✓ |
| Participant feedback questionnaire – Intervention group only |  | ✓ | ✓ |  |  |

<sup>a</sup> Conducted in-person,<sup>b</sup> Conducted via telephone
<sup>63</sup>*Revised sample size:* At the mid-recruitment DSMC review (blinded to treatment effects), observed attrition was **9%** versus the planned 20%. The DSMC recommended modifying the total sample size to **n=320**, taking into account the observed attrition rate.

Randomisation will be conducted in REDCap electronic data capture tool hosted at the Auckland University of Technology^50,51^ to either HWC or UC by the data manager (NH). Stratified randomisation will be used to balance prognostic factors via a minimization algorithm. The strata will be constructed using the combination of age (2) [<55 vs ≥55 years], sex (2) (female vs male), and ethnicity (4) [European, Pacific, Māori, Asian and other). Allocation will be concealed within REDCap; only the study managers and data manager will have access to the randomisation module and will trigger randomisation after eligibility is confirmed.

### The HWC intervention

Health coaches delivering the intervention receive training from an Internal Coaching Federation (ICF) approved trainer. The HWC intervention will be based core coaching competencies and code of ethics, developed by the International Coach Federation (ICF) to support greater understanding of the skills and approaches used in the coaching profession.^1,2^. ICF coaching is an internationally recognized approach effectively used in various settings.^3^ Following randomisation, participants will be informed of their allocation to the HWC or UC group by the study coordinator/manager. Those who are in the HWC group will be assigned a health coach who had undergone study specific health coaching training. The coaching session content and materials are described in Supplementary Table 1. Health coaches will rate the coachability of participants as described in Supplementary Table 2. Coaching sessions will be conducted in-person initially (the first 1-2 sessions) followed by telephone or video conference sessions, with the goal of up to 12 sessions in total. The timing and frequency of the sessions will be weekly, fortnightly, then monthly, but may be tailored to meet the needs of the participants (details are described in Supplementary Table 3.1 to 3.8). Individual allocated to the HWC group will also be encouraged to use the validated and free to use Stroke Riskometer app, (a mobile app that provides information about stroke, allows individuals to calculate their risk of stroke, information to support the management their risk factors)^52,53^ endorsed by the World Stroke Organization, World Heart Federation, and European Stroke Association. All coaching sessions will be audio recorded for subsequent fidelity checks. The number of coaching sessions received will be recorded in the study database.

### Primary Outcome

The **primary objective** is to determine the effectiveness of HWC in improving blood pressure at 6 months post-randomisation.

### Secondary Outcomes

The **secondary objectives** are to determine the effectiveness of HWC in the improving cardiovascular health (measured by LS7) at 6-months post-randomisation and longer term (i.e., 12-month), improving awareness about stroke symptoms, risk factors and their management 6- and 12-months post-randomisation compared to baseline, changes in cognitive functioning (as measured by the Montreal Cognitive Assessment (MoCA)),^54^ mood (as measured by Hospital Anxiety and Depression Scale),^55,56^ life satisfaction (as measured by the Satisfaction with Life Scale),^57,58^ 5-year stroke risk (as measured by the Stroke Riskometer app),^59^ health care cost, productivity level (as measured by the Consolidated health economic evaluation reporting standards (CHEERS) statement,^60^ medication adherence usings the self-efficacy for appropriate medication use scale (SEAMS) questionnaire^61^ and participants’ quality of life using the EuroQol 5D^62^ and new CVD (including stroke) events and hospitalisations. We will also examine these outcomes in individuals randomised to HWC who used and did not use the Stroke Riskometer app.

Follow-up assessments will be conducted at 3-, 9- and 12-months by telephone and in-person at 6- months post-randomisation. Blood glucose and blood cholesterol will be measured using a validated point of care blood test (CardiChek PA Analyser, https://www.pocd.co.nz/products/cardiochek-pa-analyser).

Outcome assessments will be conducted as outlined in Table 1.

### Coaching compliance and coaching evaluation

*Compliance* with health coaching will be assessed as session attendance. Sessions will be recorded as completed or missed, with reasons for missed sessions recorded. Coaching attendance will be categorised as: 0-2 no coaching; 3 - 5 some coaching and 6-12 full coaching.

Health coaches will self-evaluate each session using the *Coaching Evaluation* questionnaire. In addition, 5% of recordings will be evaluated by the coach trainer and supervisor to track the quality of coaching and identify any potential areas of improvement. This will be used to guide ongoing supervision of the coaches. These trainer/supervisor evaluations will be used to indicate reliability of the coaching evaluations.

Participant coachability will be subjectively assessed by health coaches as; (1) highly coachable; (2) quite coachable; (3) moderately coachable, and (4) not coachable. See supplemental Table 2 for coachability criteria.

### Data safety monitoring Committee

An independent Data Safety Monitoring Committee (DSMC) will oversee the trial. The DSCM will include an independent statistician, an independent stroke expert, and a community representative. A study DSMC charter approved by the committee will be used as the guiding document. The DSMC chair will review study reports to advise on any issues regarding participant safety, efficacy of the trial, including recruitment and study design parameters, and adherence to the study protocol. The DSMC will meet twice a year or as required through the duration of the trial. Interim analyses may be conducted when 50% of participants have been recruited, if advised by the DSMC.

## Statistical analysis plan

The primary analyses will be conducted using the principle of intention to treat (ITT) by a statistician blinded to group randomization with only labels (A/B) until primary results are finalized. All randomised participants who contributed at least one measure of an outcome will be included in the ITT analysis according to the group to which they are randomised. Per-protocol analysis will be used as the sensitivity analysis.

### Statistical analysis plan

All analyses will follow a pre-specified statistical analysis plan and be conducted according to the intention-to-treat principle. Descriptive statistics will summarise baseline characteristics and follow-up measures overall and by study group. Continuous variables will be reported as means (95% confidence intervals) and standard deviations or medians and interquartile ranges, as appropriate. Categorical variables will be summarised using frequencies and percentages. Analyses will be two-sided with a significance level of p < 0.05, unless otherwise stated.

The primary outcome (change in systolic blood pressure) and most secondary outcomes (e.g. Life’s Simple 7 score, Montreal Cognitive Assessment, EQ-5D-5L, SEAMS, life satisfaction, mood scores) are continuous and measured repeatedly over time (baseline, 3, 6, 9 and 12 months). Secondary analysis details are described in Supplementary Table 4.

Linear mixed-effects models (LMMs) will be used as the primary analytical approach to model longitudinal changes in outcomes and to compare trajectories between the Health Wellness Coaching (HWC) and usual care groups. These models appropriately account for within-participant correlation arising from repeated measurements and allow inclusion of participants with incomplete follow-up under a missing-at-random assumption.^4^

Models will include fixed effects for treatment group, time, and group-by-time interaction, with adjustment for baseline outcome values, stratification factors (age, sex, ethnicity), recruitment site, and relevant clinical covariates (e.g. comorbidities, baseline LS7, medication adherence). A random intercept for participants will be included, with additional random effects for site or health coach if clustering is detected.

Time will be treated as a categorical variable to allow flexible estimation of treatment effects at each follow-up point. Linear, quadratic or higher-order trends will be explored in sensitivity analyses. Model assumptions will be assessed using standard diagnostic procedures. If outcome distributions substantially deviate from normality, appropriate transformations will be considered. Where transformation is not suitable, mixed-effects quantile regression will be used to estimate median differences between groups while accounting for clustering and repeated measurements.

Time-to-event outcomes, including incident cardiovascular disease (CVD), stroke events, and hospitalisations, will be analysed using competing-risk regression methods to account for death as a competing event. Mutually exclusive outcome frameworks will be applied^4^, including: (a) death versus non-fatal CVD events; (b) fatal versus non-fatal stroke; and (c) stroke versus transient ischaemic attack. Cause-specific hazard models will be selected as appropriate. Covariate adjustment will mirror the primary analysis, with stratification by recruitment site if required.

Among participants randomised to HWC, dose–response analyses will examine the association between programme adherence (number of coaching sessions attended) and primary and secondary outcomes. Coaching quality indicators (e.g. coach ratings, trainer assessments) will be explored as potential confounders or effect modifiers. Linear or non-linear mixed models will be used, with quantile regression applied if outcome distributions are non-normal. Secondary exploratory analyses will compare outcomes among HWC participants who used versus did not use the Stroke Riskometer app. Sensitivity analysis for Intervention Compliance will include the number of coaching sessions completed, coaches rating of participant coachability, the coaching trainer’s rating of 5% of the coaching interviews conducted and the average of coaches’ rating score for each session.

Missing outcome data will be handled under the missing-at-random assumption using multiple imputation by chained equations. Imputation models will include treatment group, baseline outcome values, demographic variables (age, sex, ethnicity), and relevant clinical predictors (e.g. comorbidities). Results will be pooled across imputed datasets using Rubin’s rules.^5^

Adjustment for multiple comparisons will be applied for secondary outcomes using the Benjamini– Hochberg false discovery rate approach.^6^ Sensitivity analyses will assess robustness of findings to assumptions about missing data, model specification, and intervention compliance, including measures of coaching exposure and quality.^7^

6465666567

### Adherence and protocol deviations

Protocol deviations and violations and their reasons will be recorded by the study manager throughout the trial. Adherence to the intervention in terms of the number of coaching sessions attended and the quality of the coaching (scored by the health coach after each session) will be recorded in the e-CRFs.

### Severe adverse outcomes

The Severe Adverse Events form will be completed whenever a participant has a known hospital admission and/or by cross-checking medical records at 6- and 12-months follow-up periods. Events will be coded into stroke, TIA, myocardial infarction, or other. The SAE and interventional-related AE will be compared using chi-squared/Fisher Exact tests between the two groups.

## Discussion

The TIIPS trial will provide evidence of the efficacy of HWC for secondary stroke prevention. It will also provide guidance on tailoring the coaching intervention for stroke and TIA patients, specific to their needs compared to the general population. The effectiveness of the intervention and learnings from tailoring the intervention for the stroke population could lead to its implementation in clinical practice, resulting in clinically meaningful reduction in stroke risk.

## Data Availability

No datasets were generated or analysed for this study. This manuscript describes a study protocol; therefore, data sharing is not applicable. Any data used in the future study will be obtained from published sources or made available from the corresponding author upon reasonable request, subject to relevant data access and ethical approvals.

